# Shared genetic architecture between anorexia nervosa and metabolomic biomarkers suggest underlying causal pathways

**DOI:** 10.64898/2025.11.29.25341234

**Authors:** Carolina Makowski, Alexey Shadrin, Sara E. Stinson, Nora R. Bakken, Helga Ask, Alexandra Havdahl, Anders M. Dale, Ole A. Andreassen, Dennis van der Meer

## Abstract

**Background:** Anorexia Nervosa (AN) has a high mortality rate and often a chronic illness course, but lacks effective treatments. AN is heritable and shares genetic architecture with cardiometabolic traits, while the relationship to metabolomic markers is unknown.

**Methods:** We examined shared genetic architecture between AN and 249 metabolomic biomarkers, and compared profiles with related mental health, anthropometric and cardiometabolic traits. Genetic and biological overlap was assessed with global genetic correlations, conjunctional false discovery rate, bivariate Gaussian mixture modeling, and gene set enrichment analysis across body tissues. We also explored causal relationships between AN, body mass index (BMI), and metabolomic biomarkers.

**Results:** Significant genetic correlations were found between AN and 142 metabolomic biomarkers, which were opposite in direction to cardiometabolic traits such as BMI and type 2 diabetes (r_s_<-0.94), and stronger than correlations with anxiety. Shared variants between AN and metabolomic biomarkers, particularly lipid-based metabolites, exhibited a mixture of positive and negative effects. These were mapped to genes involved in lipid-related cell signals, developmental growth, and inflammation, and were widely expressed in the brain, most internal organs and female reproductive organs. Causal bidirectional influences between AN and metabolomic biomarkers were found to act through their respective influence on BMI.

**Conclusions:** Strong associations between AN and metabolomic markers, opposite in direction to anthropometric and cardiometabolic traits, are driven by developmental and lipid-based biological processes, with a potential mediating role of BMI. The findings offer a novel perspective on the mechanisms of AN and suggest opportunities for targeting specific metabolomic biomarkers in weight restoration approaches.

## INTRODUCTION

Anorexia nervosa (AN) is a severe psychiatric disorder marked by extreme food restriction, an intense fear of weight gain, and a distorted perception of body image. AN has a strong cardiometabolic component, with emerging evidence from both genetic and phenotypic levels of investigation urging the field to revisit this aspect of the disorder (1–4). However, there is a need to elucidate the relationship between AN and disrupted systemic metabolism (e.g., through circulating metabolomic biomarkers) that cannot be captured with cardiometabolic traits alone. Given limited treatment efficacy and high mortality rates (5–7), a better understanding of the shared biological mechanisms between AN and metabolic markers may offer new insights into the etiology of this complex disorder.

Genetic studies indicate that AN has a substantial heritable component, with twin-based heritability estimates ranging from 48-74%, underscoring the importance of genetic factors in its etiology. AN is often comorbid with other psychiatric conditions, particularly anxiety (8,9), with two-thirds of individuals with AN reporting one or more lifetime anxiety disorders (8). Recent efforts to map the genetic architecture of AN through genome-wide association studies (GWAS) have highlighted significant overlap not only with other psychiatric traits, but also with cardiometabolic traits, including anthropometric measures (body mass index [BMI]), insulin sensitivity, glycemic-, and lipid-related traits (1,10–12), and shared risk variants with metabolic pathways (13). Gut hormone levels that are fundamental to systemic metabolism are altered in patients with AN, such as increased ghrelin and peptide YY, and decreased glucagon-like peptide-1 (GLP-1) and leptin levels (4,14–17). Longstanding clinical observations have also underscored the impact of metabolic factors on weight restoration in patients, which are essential for both somatic and mental health recovery. Altogether, this has led to a re-conceptualization of AN as a metabo-psychiatric disorder (4).

BMI in particular has garnered attention in understanding the genetically-informed etiology of AN (18). A strong relationship exists between these two traits both phenotypically (e.g., low BMI is a diagnostic criterion for AN) and genetically, with high genetic correlation and putative bi-directional causal associations (1). Importantly, previous GWAS have found that many of the discovered genetic variants associated with AN are also independent of the effects of BMI (1). It is still unclear how the biological factors shaping BMI may increase risk for AN, and how AN’s impact on BMI may influence systemic metabolism. Recent genetically-driven evidence indicates that BMI causally influences circulating metabolite concentrations, with biological processes related to lipid homeostasis (19). It is now possible to determine the role of circulating metabolomic biomarkers in relation to BMI with high-throughput screening technology applied in large-scale biobanks (20). Understanding these relationships is imperative to improve weight restoration during AN treatment (18).

Circulating metabolomic biomarkers represent a novel opportunity to better understand the biological pathways underlying the close relationship between anthropometric, cardiometabolic factors, and AN (21). These metabolomic biomarkers encompass lipoproteins, ketone bodies, fatty acids and amino acids that are key markers of systemic metabolism and indicators and predictors of human health (19,22). While prior work has elucidated key links between AN and broad anthropometric and cardiometabolic traits, the relationship between AN and individual circulating metabolomic biomarkers is unknown. Altered metabolomic marker levels have been reported in children (7 years old) prior to an AN diagnosis in adolescence (14-18 years), particularly in high- and very-low density lipoproteins (HDLs, VLDLs), docosahexaenoic acid, poly- and mono-unsaturated fatty acids, and triglycerides (2), providing evidence for a biological predisposition to AN. A better understanding of the shared biology between AN and circulating metabolomic biomarkers could inform treatment targets in patients with AN. Assessing metabolite levels as a measure of treatment efficacy are engrained into medical care and could provide powerful new leads for the treatment of AN.

Here, we examine the shared genetic architecture between AN and metabolomic markers by leveraging GWASs of AN and 249 circulating metabolomic biomarkers. We compare global genetic overlap between metabolomic markers and AN to commonly comorbid psychiatric conditions (e.g., anxiety) and anthropometric and cardiometabolic traits (e.g., BMI, type 2 diabetes, coronary artery disease). We leveraged cross-trait genetics to identify shared genetic variants between AN and circulating metabolomic biomarkers to pinpoint shared underlying biological pathways. Finally, we aimed to better understand the putative bidirectional causal relationships between BMI and AN, through integration of genetic risk of circulating metabolomic biomarkers in mendelian randomization analyses. This approach could offer fruitful avenues for future investigations and development of novel treatment alternatives.

## METHODS AND MATERIALS

### Metabolomic markers & GWAS summary statistics

Metabolomic markers consisted of 249 circulating metabolomic biomarkers generated using Nightingale Health’s high-throughput Nuclear Magnetic Resonance (NMR) metabolomics platform (20), processed and QC’ed using the ‘ukbnmr’ pipeline. See Supplementary Table 1 for a full list and Supplementary Material for more information on these markers. Summary statistics were drawn from separate GWAS run on these 249 metabolomic biomarkers (19). The discovery GWAS of metabolomic markers included a meta-analysis across 207,841 participants from the UK Biobank and 92,661 participants from the Estonian biobank (19). We also conducted a replication GWAS of metabolomic markers derived from a hold-out sample of 166,530 participants from the UK Biobank (see Supplementary Materials).

#### GWAS summary statistics: Psychiatric, anthropometric and cardiometabolic disease traits

Summary statistics for AN were taken from the latest published AN GWAS of 16,992 individuals with AN and 55,525 controls (1). Additional summary statistics were included for anthropometric (BMI (23)), cardiometabolic (Type 2 Diabetes (T2D) (24), and Coronary Artery Disease (CAD) (25)), and psychiatric (Anxiety (ANX) (26)) traits. See additional details in Supplementary Table 2. All summary statistics were processed through a standardized pipeline to ensure consistency (https://github.com/precimed/python_convert/blob/master/sumstats.py). We excluded all variants within the extended major histocompatibility complex (MHC) region (chr6:25–35 Mb).

### Bivariate LDSC

We applied cross-trait LD score regression (LDSC; v1.0.0; https://github.com/bulik/ldsc) (27) to estimate genetic correlations between metabolomic markers, AN, ANX, BMI, T2D, and CAD. Correction for multiple comparisons across the 249 circulating metabolomic markers were carried out using the Benjamini and Hochberg method, with adjusted significance thresholds set at α*=*.*05*.

### Conjunctional FDR to assess AN-metabolomics overlap

We applied a conditional/conjunctional false discovery rate (condFDR/conjFDR) framework with default settings (https://github.com/precimed/pleiofdr) to identify shared genetic variants between AN and 249 metabolomic markers. See Supplementary Material for more details. This framework is a model-free strategy to assess summary statistics from GWAS and can capture both agonistic and antagonistic directional effects of genetic variants associated with two traits (28–30).

### Bivariate MiXeR

To estimate the proportion of shared causal genetic variants between AN and metabolomic markers, we applied a bivariate Gaussian mixture model, MiXeR (31,32). See Supplementary Materials for more details. We prioritized metabolomic markers that had a strong positive or negative genetic correlation relative to other markers with AN as determined through LDSC, as well as a high number of overlapping genetic variants, relative to other metabolites, as defined by conjFDR analyses.

### Gene Mapping of overlapping AN-metabolomics variants

We used the Variant-to-Gene pipeline from Open Targets Genetics to map lead variants from the above-mentioned cFDR analyses to genes. This pipeline maps genes based on the strongest evidence obtained from quantitative trait loci (QTL) experiments, chromatin interaction experiments, in silico functional prediction, and proximity of each variant to the canonical transcription start site of genes (33).

### Gene set enrichment analyses

We carried out gene set enrichment analyses on the mapped genes, investigating terms that are part of the Gene Ontology biological processes subset (n=7522), as listed in the Molecular Signatures Database (MsigdB; c5.bp.v7.1). We ran Fisher exact tests on the 249 sets of mapped genes between each metabolomic marker and AN, leveraging the enrichR R package. We restricted the output to those terms that had more than 4 overlapping genes and that were smaller than 1000 genes in total, with a q-value <.05. We further removed terms from the list of significant enrichments if more than 80% of their overlapping genes were among other more significant terms.

### Tissue enrichment

We leveraged tissue-specific gene expression databases to better understand the gene expression patterns of uncovered shared genetic variants and their mapped genes to different organs. Specifically, we investigated the overlap between the above-mentioned mapped genes from cFDR analysis across all AN-metabolic marker pairs, and differentially expressed genes (DEGs) in 30 general tissue types available in the GTEXv8 database. See Supplementary Materials for details. Given the strong neurobiological underpinnings of AN, we further investigated overlap between our mapped variants/genes and brain-based gene expression by incorporating mRNA distribution data from the Allen Human Brain Atlas (34). Brain data were summarized with the Desikan-Killiany parcellation and mapped with the Python toolbox Abagen (35). Data normalization was performed using the default scaled robust sigmoid method (36). To assess the difference in gene expression between the list of identified genes and all other genes, a Wilcoxon rank-sum test was conducted for each brain region.

### Mendelian randomization

We ran bidirectional Mendelian Randomization (MR), investigating the causal relationships between AN, BMI, and the 249 circulating metabolomic markers. The TwoSampleMR R package (https://mrcieu.github.io/TwoSampleMR/) (37) was applied to GWAS summary statistics. We selected only genome-wide significant variants for the analysis, clumped using PLINK with clump_p=1, clump_r2=0.001, clump_kb=10000 against the 1000 Genomes Phase3 503 EUR samples, keeping other settings default. We calculated MR regression coefficients using the inverse variance weighted (IVW) method and further applied the weighted median method to ensure robustness of findings (38), where significance was defined as q<0.05 based on FDR correction for both methods.

## RESULTS

### Genetic correlations between AN and metabolomic markers

Global genetic correlations, assessed with LDSC, showed significant genetic correlations between AN and 142 of the 249 metabolomic biomarkers after correction for multiple comparisons (max |r_g_|=0.35, p-adjusted<0.05) (Figure 1A and B). Genetic correlation patterns were largely similar when comparing AN and other traits to a metabolomic GWAS applied to a hold-out replication dataset from the UK Biobank, where 161 of the 249 metabolomic markers had significant genetic correlations with AN (Supplementary Figure 1, Supplementary Table 3). Genetic overlap was highest for lipoprotein subclasses, relative lipoprotein lipid concentrations and amino acids, with a relatively equal number of significant positive (68) and negative (74) genetic correlations (Supplementary Table 3). Genetic correlations between AN and metabolomic markers were generally opposite in direction to associations found with anthropometric and cardiometabolic disease traits, and anxiety (Figure 1B; Supplementary Figure 1B). Correlation strengths with the AN-metabolomic marker profile were strongest with the cardiometabolic disease traits (r_s_=−0.91 to −0.95), and negatively correlated with anxiety (r_s_=−0.89) (Figure 1C). Some metabolomic associations emerged with AN that were not found for anxiety, particularly for branched-chain amino acids (e.g., leucine, valine) which showed significant negative genetic correlations with AN. Spearman’s correlations between metabolomic genetic correlation profiles with individual traits were stronger than the genetic correlation strengths between the traits themselves (Figure 1C; Supplementary Figure 1C), and even in the opposite direction as was the case for AN and anxiety (positive r_g_=0.35, p=0.034).

**Figure 1.**
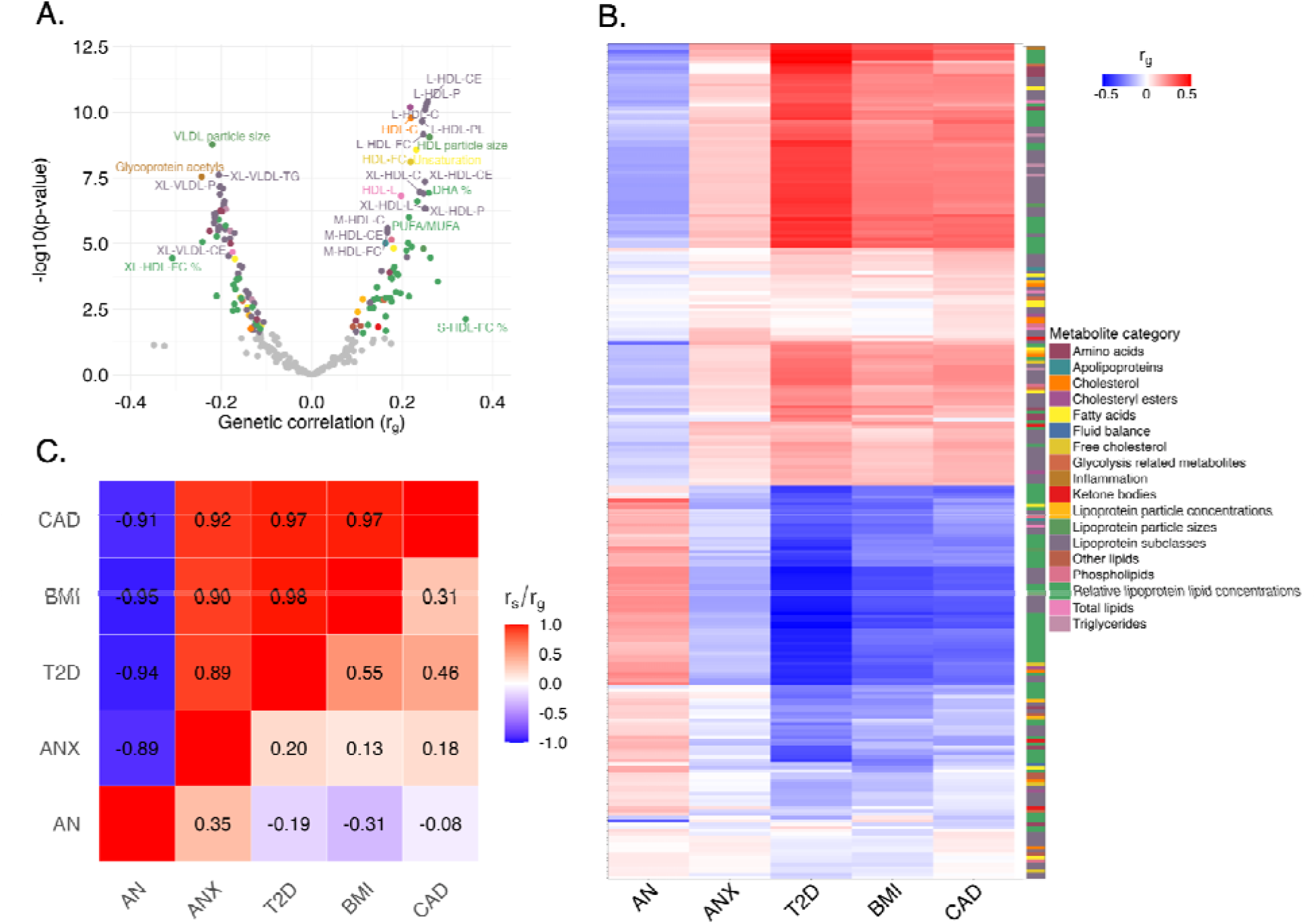
Genetic correlations between metabolomic markers and AN, and comparison to other traits. A: Volcano plot summarizing genetic correlations between AN and all 249 circulating metabolomic biomarkers, with significant correlations depicted in color, corresponding to the legend in panel B. See Supplementary Table 1 for full names of selected annotated markers. B: Heatmap of genetic correlation profiles of AN and four other psychiatric, anthropometric, and cardiometabolic traits (x-axis), with the 249 metabolomic markers along the y-axis. The y-axis is sorted by hierarchical clustering, with the legend indicating metabolic categories. C: The upper triangle of the correlation matrix depicts Spearman’s correlations of genetic correlation profiles (shown in Panel B) between the five traits and metabolomic markers. The lower triangle depicts genetic correlations between the five traits. Abbreviations: AN. Anorexia Nervosa; ANX. Anxiety; T2D. Type 2 Diabetes; BMI. Body Mass Index; CAD. Coronary Artery Disease; r_g_. genetic correlation; r_s_. Spearman’s correlation.

### Shared genetic variants between AN and metabolomic markers

We leveraged conjunctional FDR to identify specific genetic variants shared between AN and metabolomic markers for further downstream biological interpretation. The majority of metabolomic markers (240 of the 249) had at least one associated variant shared with AN, with up to 16 shared variants for two of the metabolomic markers (Figure 2A, Supplementary Table 4). These two markers were cholesteryl esters in HDLs (HDL-CE) and triglycerides to total lipids in small HDL percentage (S-HDL-TG pct). Thus, leveraging a cross-trait genetic overlap approach effectively doubled the number of discovered variants compared to the 8 variants uncovered in the AN GWAS (1).

**Figure 2.**
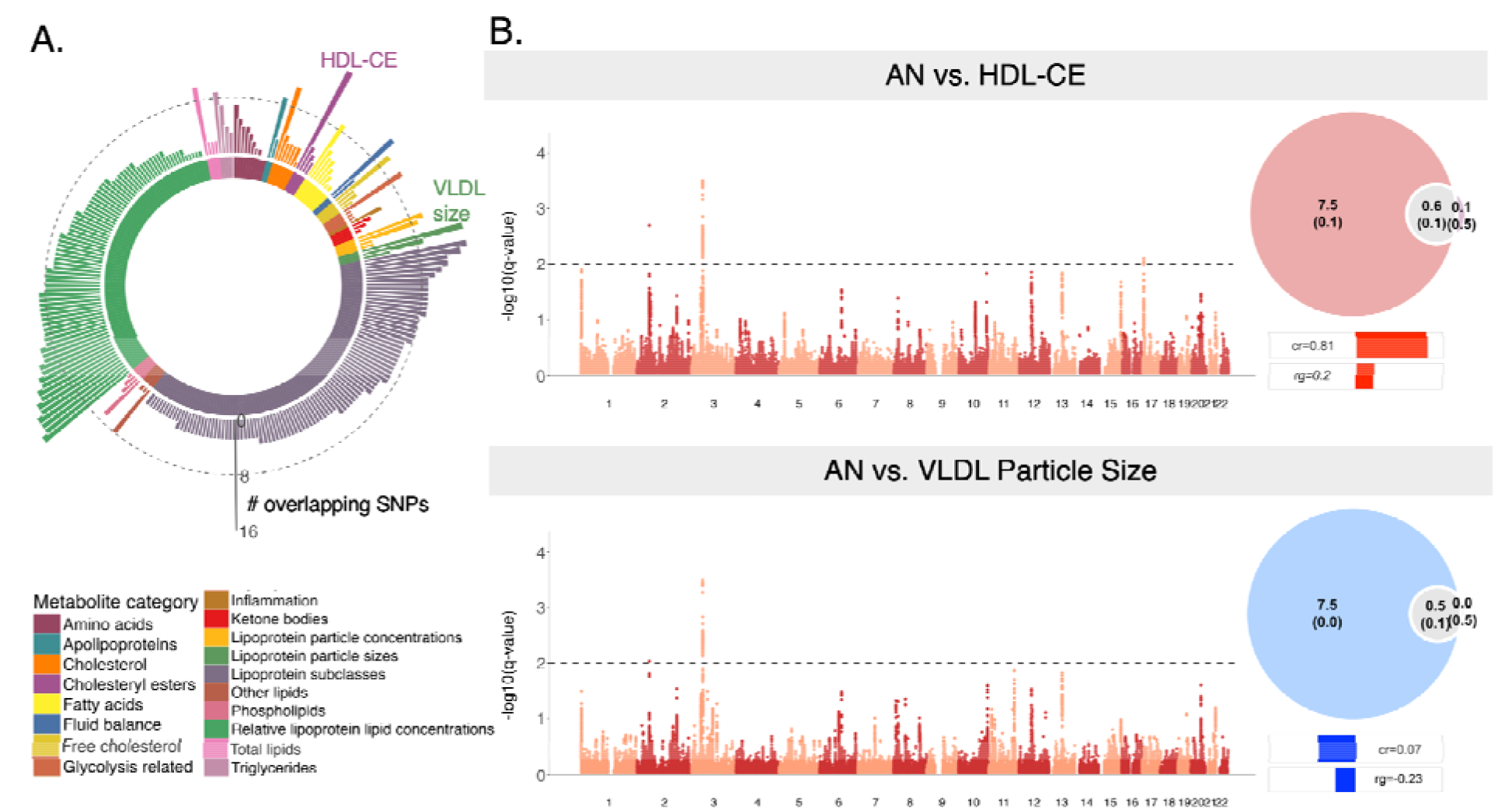
Degree of genetic overlap between AN and metabolomic markers. A. Circos plot depicting the number of significant genetic variants or SNPs overlapping between AN and each of the 249 circulating metabolomic biomarkers determined through conditional/conjunctional FDR analysis, with the height of the bar corresponding to the number of overlapping variants ranging from 0-16, colored by metabolic marker. The dotted line reflects 8 overlapping SNPs, denoting the number of genome-wide significant SNPs found in the original AN GWAS, whereby cFDR analyses with many metabolic markers significantly boosted discovery of genetic variants associated with AN. Two selected metabolic markers are labeled in A and further depicted in panel B: Manhattan plots of genome-wide significant SNPs associated with both AN and 2 select metabolic markers, HDL-CE and VLDL Particle Size, with 16 and 15 overlapping SNPs, respectively. Accompanying Venn Diagrams illustrate the genetic overlap between AN and these two markers after Bivariate mixture modeling. The estimated number of unique and shared variants are indicated in thousands, with standard deviations in brackets. Bars below the Venn diagrams indicate the concordance rates (“cr”) in direction of effects on the pair of traits, with their genetic correlation (“rg”) listed as a reference. Abbreviations: AN. Anorexia Nervosa; SNPs. Single Nucleotide Polymorphisms; HDL-CE. Cholesteryl esters in high-density lipoproteins; VLDL. Very-low-density lipoproteins.

A larger proportion of shared variants (57.9%) had discordant effects (i.e. acting in opposite directions) between AN and metabolomic markers than concordant effects (□^2^(1)=31.47, p=2.02e-8) (Supplementary Table 5). Given this mix of direction of effects, alongside the polygenic nature of these traits (39), we leveraged bivariate Gaussian mixture modeling with MiXeR (31) to quantify the extent of genetic overlap between AN and two select metabolic markers regardless of effect directions: HDL-CE and diameter or particle size of VLDLs (Figure 2B, Figure 2C). These markers were prioritized given their strong genetic correlations with AN, both positive (HDL-CE: r_g_=0.22, p=6.37e-11) and negative (VLDL Particle Size: r_g_=−0.22, p=1.7e-9), with a high number of overlapping genetic variants as determined by cFDR (16 and 15 overlapping variants for HDL-CE and VLDL Particle Size, respectively) (Figure 2B). Analysis with MiXeR revealed substantial genetic overlap for both HDL-CE and VLDL particle size (Figure 2C) (Supplementary Table 6). Concordance rates with MiXeR demonstrated a mixture of positive and negative effects, where 81% and 7% of the shared variants were estimated to have concordant effects between AN and HDL-CE/VLDL Particle Size, respectively (Figure 2C).

### Shared biological pathways and tissue enrichment

We assigned the uncovered shared genetic variants between AN and metabolomic markers to genes using Open Targets Genetics (Supplementary Table 5). Across all AN-metabolomics pairs, 80 unique genes were found. These genes were tested for enrichment of biological processes using Gene Ontology (GO) analyses (Figure 3A, Supplementary Table 7). We found 13 significant GO terms of shared biological processes across AN and 7 metabolomic markers. Notable pathways involved biological processes related to lipid-related cell signals, developmental growth (e.g., increase in size or mass of an organism or cell), inflammation (e.g., cytokine production) and canonical cellular processes (e.g., tyrosine kinase signaling) (q<0.05) (Figure 3A).

**Figure 3.**
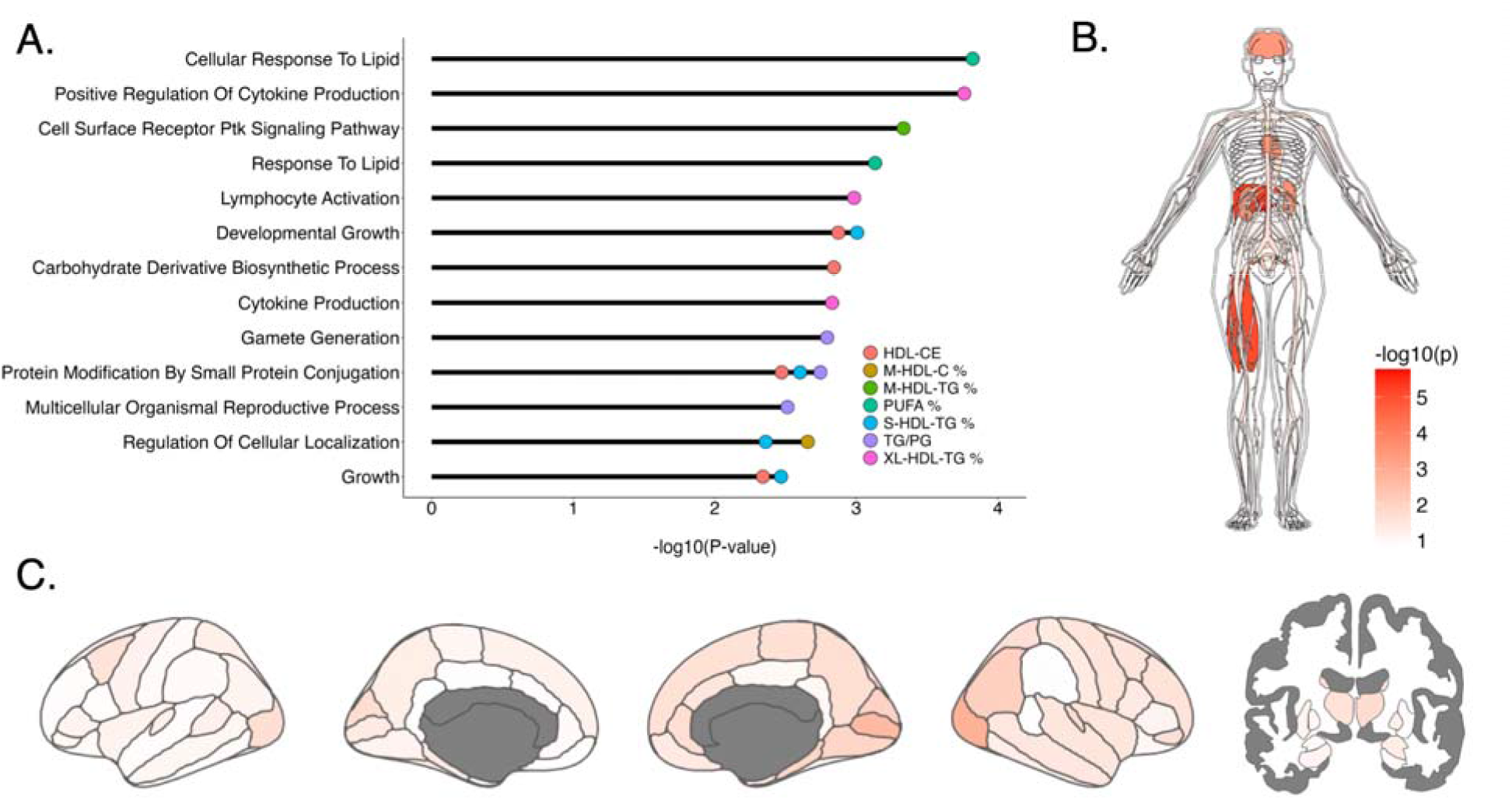
Biological pathways and tissue-specific gene expression enrichment of shared AN-metabolomic genes. A. Significantly enriched gene ontology GO terms for mapped shared genes between AN and metabolomic markers, indicated by different colors. B. Anatogram depicting enrichment of shared genes between AN and metabolomic markers across 20 different tissue types from the GTEx database. C: Brain maps showing statistics of enrichment tests across cortical (Desikan-Killiany atlas) and subcortical (Freesurfer aseg atlas) regions, based on expression data from the Allen Human Brain Atlas; note no brain regions survive correction for multiple comparisons (where nominal p=0.05 corresponds to −log10(p)=1.30). Panels B and C depict −log10(p)-values on the same scale, as indicated by the color bar.

We then investigated the gene expression patterns of mapped genes across the brain and other organs. We found significant enrichment of overlapping AN-metabolomic genes and gene expression patterns across 10 of the 20 examined organs, including brain, pancreas, liver, heart, kidneys, skeletal muscle, leukocyte and female reproductive organs (q<0.05) (Figure 3B) (Supplementary Table 8). Nominally significant associations emerged with enrichment for gene expression of overlapping AN-metabolomic marker genes in right lateral occipital and pericalcarine regions (unadjusted p<0.05), but did not survive correction for multiple comparisons (Figure 3B; Supplementary Table 9).

### Causal relationships between AN, BMI and metabolomic markers

We investigated potential causal associations between AN and metabolomic markers using Mendelian randomization (MR), and potential interrelationships with BMI, given its inherent phenotypic relationship with AN and strong negative genetic correlation (r_g_=−0.31) compared to other cardiometabolic disease traits (r_g_<-0.19 for T2D and CAD). No evidence was found for a causal association between AN and metabolomic markers in either direction. However bi-directional associations were found between AN and BMI (Figure 4A) (Supplementary Table 10), confirming previously reported bi-directional relationships (1), with a stronger effect size for the presence of AN putatively causing variation in BMI (IWV method, β=−0.44, *p*=1.06e-19) compared to BMI causing AN (β=−0.06, *p*=4.26e-4). Significant bi-directional associations were also found between BMI and various metabolomic markers (Figure 4A) (Supplementary Tables 11, 12), although the majority of effects suggested a causal direction from BMI to metabolomic markers, with 6 markers showing both a putative causal impact on BMI, as well as being shaped by BMI (Figure 4B).

**Figure 4.**
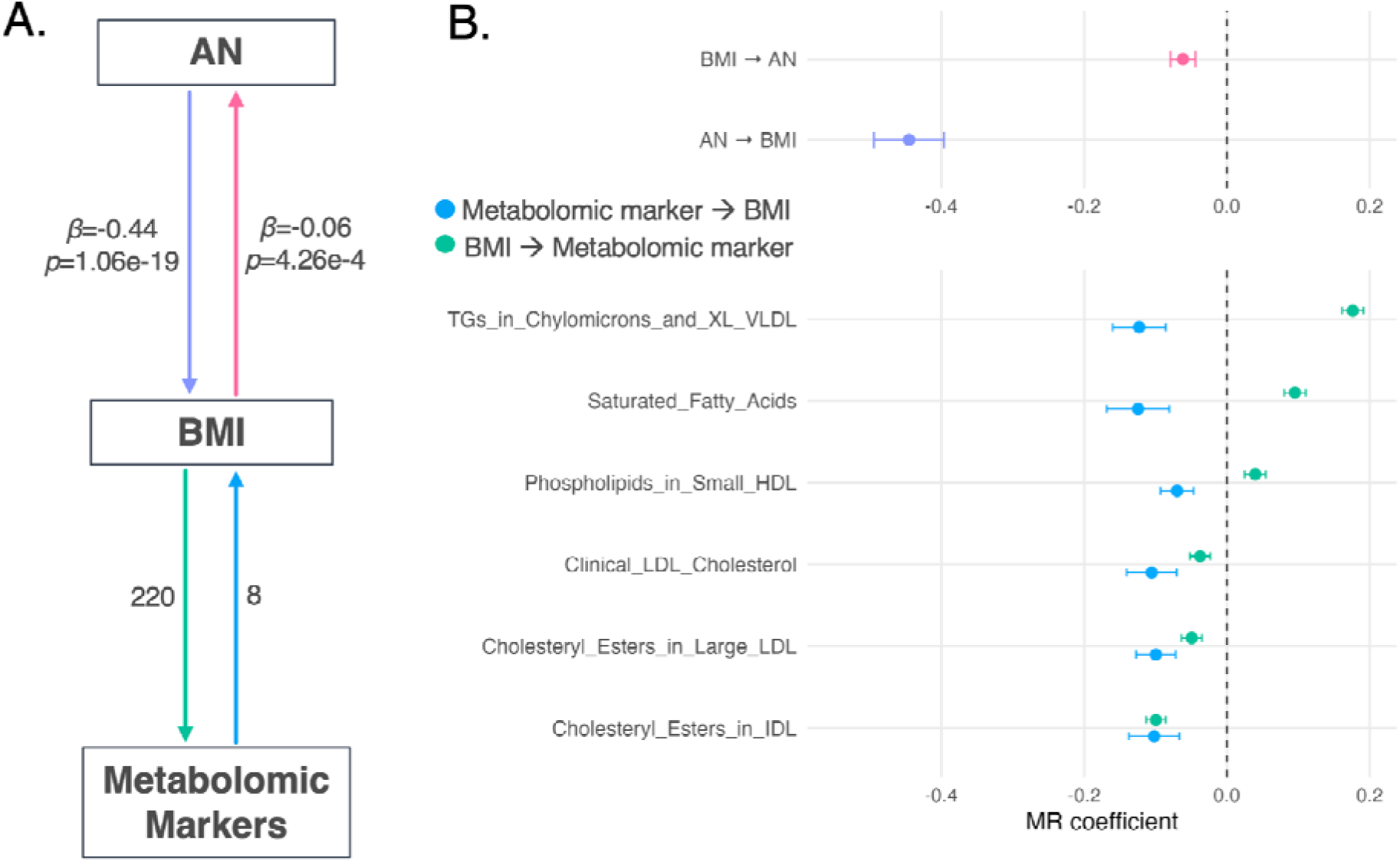
Bidirectional causal relationships between AN, BMI, and metabolomic markers. A: Schematic summarizing results from Mendelian Randomization (MR) analyses, whereby AN and metabolomic markers may be linked indirectly through shared causal mechanisms with BMI. Top half of panel A includes labels with statistics derived from the inverse weighted variance method, and labels next to arrows on the bottom half indicate the number of metabolic markers that contribute to the putative causal relationship between BMI ←→ Metabolomic markers. B: MR coefficients derived from the inverse weighted variance method depicted for both BMI ←→ AN, and the six metabolomic markers that bidirectionally associated with BMI. Abbreviations: AN. Anorexia Nervosa; BMI. Body Mass Index; TGs. Triglycerides; XL. Extra Large; VLDL. Very-low-density lipoprotein; HDL. High-density lipoprotein; LDL. Low-density lipoprotein; IDL. Intermediate-density lipoprotein.

## DISCUSSION

This work demonstrated strong genetic overlap and identified key genes and biological pathways shared across AN and metabolomic markers. The observed associations between AN and metabolomic markers showed a striking pattern that was similar in strength but opposite in direction compared to anthropometric and cardiometabolic disease traits. Our results suggest that developmental and lipid-based biological processes, as well as a mediating role of BMI, are key to the relationship between AN and disrupted systemic metabolism.

Our results from conjunctional FDR and bivariate gaussian mixture modeling mapped the shared genetic architecture between AN and metabolomic markers in finer detail, and uncovered both positive and negative associations between AN and various metabolomic markers. This mixture of effects would otherwise be obscured with global genetic correlations with broader metabolic traits. Our cross-trait genetics approach boosted the number of genome-wide significant loci associated with AN, up to two-fold when leveraging some AN-metabolic marker pairs. Data-driven gene ontology analyses provided insights into biological pathways that are shared between AN and metabolomic markers, including somatic developmental growth, inflammation and lipid-related processes, and canonical cell pathways. Shared genes between AN and metabolomic markers had notable gene expression enrichment across multiple tissues, corroborating the idea that AN’s etiology extends beyond just psychiatric and brain-based origins, but impacts many metabolically and clinically relevant organs across the body that are impacted during starvation and refeeding in patients with AN, including the pancreas, liver, ovaries, skeletal muscle, and kidneys. Finally, we showed evidence that the associations between AN and metabolomic markers may act through their influence on BMI, offering an important genetically-informed framework that could help future research better understand the biological factors contributing to fluctuations in weight restoration, a cornerstone of AN treatment.

Our results showed a unique profile of associations between AN and metabolomic markers compared to anxiety, a psychiatric disorder that is highly comorbid with AN and is positively genetically correlated, but had a negative genetic association when comparing metabolomic profiles. This suggests that these two disorders may have distinct biology through their links to systemic metabolism. In contrast to the negative associations between the AN-metabolic marker profile and cardiometabolic traits, the anxiety-metabolic marker profile showed weaker and positive genetic associations with T2D, BMI and CAD, concordant with phenotypic observations between anxiety and cardiovascular disorders (40). Several branched chain amino acids, including valine and leucine, showed a significant genetic correlation with AN but not with anxiety. These findings are intriguing as amino acid deficiencies have been reported in AN patients (41,42), as well as in animal studies of eating behaviors. For example, valine-deficient diets in mice result in the cessation of eating behaviors prior to reaching satiety (43). These amino acids also play a strong role in glucagon/GLP-1 secretion, contributing to the regulation of appetite, satiety, and lipid metabolism (44,45), and contribute to cognitive function and glutamate synthesis in the brain (46,47). Our findings offer a potential pathway from low levels of valine and leucine in AN that may reduce feelings of hunger, impacting food intake and downstream metabolic consequences and weight gain in AN specifically, which may not be observed in individuals with anxiety alone. A better understanding of an optimal balance of amino acid intake in AN could offer important insights for future treatment directions to improve weight restoration and precision nutrition efforts in patients.

Our work identified shared biological pathways between AN and metabolomic markers, which could further offer new leads for treatment targets. One lipid-based marker that consistently showed a strong genetic overlap with AN through multiple lines of evidence (global positive genetic correlation, high number of genetic overlap through conjFDR and MiXeR, shared biological pathways) was cholesteryl-ester in high-density lipoproteins, or HDL-CE. This marker shared 4 biological pathways in AN through a gene ontology analysis, highlighting pathways related to developmental growth, protein modification by small protein conjugation, and biosynthetic processes related to carbohydrate derivatives. This analysis also revealed that HDL-CE and other HDL-related metabolic markers shared several key genes with AN and multiple metabolic markers across various enriched biological annotations, notably *CTNNB1* or “*Beta-catenin”* on chromosome 3, part of the canonical *Wnt* signaling pathways important for cell differentiation and proliferation early in brain development (48,49), and has been directly implicated in neurodevelopmental disorders (49,50). *ARIH2* and *CTC1*, on chromosome 3 and 17, respectively, were also identified as shared across some HDL and triglyceride metabolic markers and AN, with roles in protein degradation processes. While these genes were identified by Watson et al., (1) in their investigation of AN genetic architecture, our results lend further support for prioritization of these genes for future biological follow-up given their links to specific metabolomic markers genetically associated with AN and their implications for understanding the biological underpinnings of the disorder. Future research would benefit from disentangling which metabolites are premorbid markers of AN vs. those that may catalyze illness progression and/or serve as prognostic indicators of AN outcomes.

Our work also aimed to understand the relationship between BMI and AN, through integration of circulating metabolomic biomarkers in mendelian randomization analyses. We did not find a significant direct causal relationship between AN and metabolomic markers, in line with previous findings with polyunsaturated fatty acids (51). However, bidirectional influences between AN and metabolomic markers may act through their respective influence on BMI, with effect sizes suggesting stronger evidence for AN causing low BMI, and in turn, altering metabolite levels. This is not surprising as BMI is an inherent part of the clinical picture of AN, both in terms of diagnosis and defining treatment milestones through weight restoration. Although concerns of the genetics of AN being confounded by BMI have been raised (18), previous and ongoing efforts to map the genetic architecture of AN have shown that the genetics of AN cannot be explained solely by the genetics of BMI (1,52). Our MR results linking AN, BMI and metabolic markers, alongside evidence from our GO analysis implicating developmental growth pathways, offer a new perspective for future work to consider the developmental trajectories and dynamic interplay between these traits to better pinpoint sensitive periods (e.g., pubertal development) that may confer greatest risk for AN (21,53).

Understanding the biological components of AN in relation to systemic metabolism could form the foundation for developing metabolic treatments. Metabolism-driven risk reduction strategies have been successfully applied in other fields of medicine; for instance, monitoring total and LDL cholesterol has found success for prediction and mitigation of coronary heart disease risk (54). A wide array of lipid-altering treatments are approved and routinely used for the treatment of cardiometabolic diseases, including lifestyle changes and medications such as statins and PCSK9 inhibitors (55). Extending these insights to AN is aligned with what has been proposed for adolescent-onset psychiatric risk more broadly (56). In our findings, negative correlations between AN and other cardiometabolic traits through their metabolomic profiles emerged, suggesting that future investigations of potential pharmacological agents for treatment of AN should include medications for cardiometabolic disease, but with opposite action of effects (e.g., lipid-increasing rather than lipid-lowering). This may also offer an important and needed line of investigation to explain why existing off-label medications for AN with lipid-altering effects, such as the antipsychotic aripiprazole, may be helping not only with psychiatric symptoms but perhaps with weight restoration and other aspects of AN treatment that have not been clearly elucidated or described to date.

This research needs to be interpreted in the context of limitations. The Nightingale panel from which the 249 metabolomic markers were derived is biased towards lipid-based markers, which may have in turn biased downstream analysis which pinpointed shared lipid-related biological processes between AN and various metabolomic markers. Certain metabolic markers, such as proteins, a more comprehensive panel of amino acids, and short chain fatty acids, would require additional metabolomics technology (e.g. mass spectrometry) that could be applied in the future. It is also unclear how well these findings would generalize to individuals with atypical AN, i.e., meeting diagnostic criteria for AN in the absence of low BMI. Caution should also be exercised when interpreting results from MR analyses, as elucidated relationships do not always point to direct causal effects.

Our findings point to strong associations between AN and metabolomic markers, opposite in direction to anthropometric and cardiometabolic traits. Our results also suggest a mediating role for BMI and implicate developmental and lipid-based homeostatic processes shared between AN and metabolomic markers. The shared biology between AN and circulating metabolomic biomarkers provides compelling evidence for investments in metabolism-informed research of this disorder to better understand the mechanisms of AN, and offer evidence-based treatments based on easily obtained and objectively measured metabolic markers.

## Data Availability

All data produced in the present work are contained in the manuscript.

https://pgc.unc.edu/for-researchers/download-results/

https://zenodo.org/records/15420219

## DATA AVAILABILITY

This study used openly available summary statistics from previously published genome-wide association studies. Anorexia nervosa summary statistics are available through the Psychiatric Genomics Consortium, located at https://pgc.unc.edu/for-researchers/download-results/. Metabolomic marker GWAS summary statistics are located at https://zenodo.org/records/15420219.

## ACKNOWLEDGMENTS/FUNDING

This work was in part performed on the TSD (Tjeneste for Sensitive Data) facilities, owned by the University of Oslo, operated and developed by the TSD service group at the University of Oslo, IT-Department (USIT). C.M. is supported by the National Institute of Mental Health (R00MH132886) and the Brain Behavior Research Foundation (#31876). D. M. is supported by the Research Council of Norway (#351751). S.E.S. is supported by the Novo Nordisk Foundation (NNF23OC0099658). N.R.B is supported by the Research Council of Norway (271555/F21). O.A.A. is supported by the Research Council of Norway (#273291, #273446, #296030, #324252, #324499, #326813), and the European Union’s Horizon 2020 research and innovation program (#847776 CoMorMent, #964874 RealMent). A.M.D. is supported by the National Institutes of Health (U24DA041123, R01AG076838, U24DA055330, OT2HL161847). H.A. and A.H. were funded by the South-Eastern Norway Regional Health Authority (#2019079).

