## Supplementary Materials for "Shared genetic architecture between anorexia nervosa and metabolomic biomarkers suggest underlying causal pathways"

*Metabolomic biomarkers.*

Metabolomic markers consisted of 249 circulating metabolomic biomarkers generated using Nightingale Health’s high-throughput Nuclear Magnetic Resonance (NMR) metabolomics platform (1). Briefly, this platform applies high-field ¹H-NMR spectroscopy to EDTA plasma samples to quantify 249 metabolic measures from a single assay. A proprietary software suite deconvolves the spectral data to determine the absolute concentrations of lipoprotein subclass particles, their lipid components, fatty acids, and various low-molecular-weight metabolites such as amino acids and glycolysis-related markers. While many of these markers represent derived ratios and thus may not be fully independent metabolic measures, we opted to include the full Nightingale panel in our analysis to maximize our ability to capture key biological processes that may not be captured by individual metabolites alone.

*GWAS samples for metabolomic markers.*

Summary statistics were drawn from separate GWAS run on the above-described 249 circulating metabolomic biomarkers (2). The discovery GWAS of metabolomic markers included a meta-analysis across 207,841 participants from the UK Biobank (mean age=57.4 years (SD = 8.0), 53.7 % female) and 92,661 participants from the Estonian biobank (mean age=50.9 years (SD = 16.2 years), 65.7% female) (2). We also included a replication GWAS of metabolomic markers derived from a hold-out sample of 166,530 participants from the UK Biobank (mean age=57.41 years (SD=7.98), 54.1% female), where GWAS was processed identically to the discovery sample.

*Conjunctional FDR to assess AN-metabolomics overlap.*

We applied a conditional/conjunctional false discovery rate (condFDR/conjFDR) framework to identify shared genetic variants between AN and 249 metabolomic markers. This framework is a model-free strategy to assess summary statistics from GWAS and can capture both agonistic and antagonistic directional effects of genetic variants associated with two traits (3–5). Notably, condFDR and conjFDR have substantially contributed to our understanding of genetic overlap between complex traits in many applications related to human health and psychology (3). First we applied CondFDR, which conditions the GWAS summary statistics for each pair of traits onto each other in both directions. We then applied conjFDR to extract the maximum of the two mutual condFDR for a specific SNP. CondFDR/conjFDR was carried out using the pleioFDR using default settings (https://github.com/precimed/pleiofdr), where we set an FDR threshold of 0.05 as whole-genome significance.

*Bivariate MiXeR.*

To estimate the proportion of shared causal genetic variants between AN and metabolomic markers, we applied a bivariate Gaussian mixture model, MiXeR (6,7). We prioritized metabolomic markers that had a strong positive or negative genetic correlation relative to other markers with AN as determined through LDSC, as well as a high number of overlapping genetic variants, relative to other metabolites, as defined by conjFDR analyses. MiXeR leverages GWAS summary statistics to model additive genetic effects as a mixture of four components: null SNPs in both traits (π_0_); SNPs with a specific effect on the first (π_1_ ) and on the second (π_2_) trait; and SNPs with a non-zero effect on both traits (π_12_). Methodological details have been described in previous work (6,7). Quality checks included inclusion of a positive AIC value for each tested model, compared to a baseline infinitesimal model, which indicates good model fit.

*Tissue enrichment.*

We leveraged tissue-specific gene expression databases to better understand the gene expression patterns of uncovered shared genetic variants and their mapped genes to different organs. Specifically, we investigated the overlap between above-mentioned mapped genes from cFDR analysis across all AN-metabolic marker pairs, and differentially expressed genes (DEGs) in 30 general tissue types available in the GTEXv8 database. DEGs are defined as genes with log2 transformed, normalized expression values (Read Per Kilobase per Million, zero-mean) with P-value ≤ 0.05 after Bonferroni correction and absolute log fold change ≥ 0.58 in a given tissue. Given the strong neurobiological underpinnings of AN, we further investigated overlap between our mapped variants/genes and brain-based gene expression by incorporating mRNA distribution data from the Allen Human Brain Atlas (8). In cases where multiple probes were available for a specific mRNA, the probe with the highest differential stability was selected (9). Brain data were summarized with the Desikan-Killiany atlas, mapped with the Python toolbox Abagen (10). Data normalization was performed using the default scaled robust sigmoid method (9). To assess the difference in gene expression between the list of identified genes and all other genes, a Wilcoxon rank-sum test was conducted for each brain region.

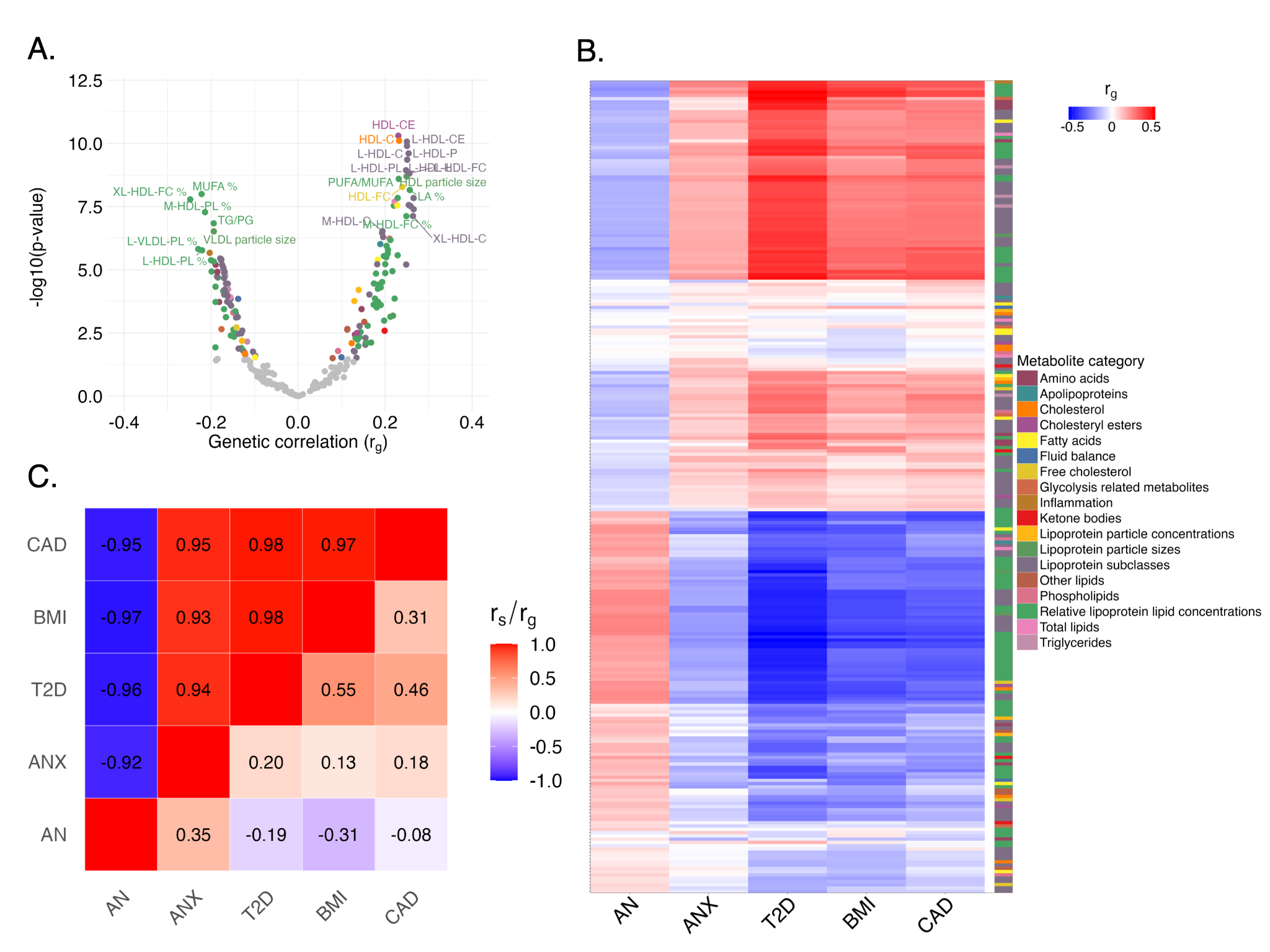

**Supplementary Figure 1**. **Genetic correlations between metabolomic markers and AN, and comparison to other traits, using an independent replication GWAS of metabolomic markers**. A: Volcano plot summarizing genetic correlations between AN and all 249 circulating metabolomic biomarkers, with significant correlations depicted in color, corresponding to the legend in panel B. Only some of the top metabolomic markers are labelled for ease of visualization. See Supplementary Table 1 for full names of selected annotated markers. B: Heatmap of genetic correlation profiles of AN and four other psychiatric, anthropometric, and cardiometabolic traits (x-axis), with the 249 metabolomic markers along the y-axis. The y-axis is ordered identically to what is presented in the main manuscript in Figure 1B, with the legend indicating metabolic categories. C: The upper triangle of the correlation matrix depicts Spearman’s correlations of genetic correlation profiles (shown in Panel B) between the five traits and metabolomic markers. The lower triangle depicts genetic correlations between the five traits, identical to what is presented in the main manuscript in Figure 1C. Abbreviations: AN. Anorexia Nervosa; ANX. Anxiety; T2D. Type 2 Diabetes; BMI. Body Mass Index; CAD. Coronary Artery Disease; r_g_. genetic correlation; r_s_. Spearman’s correlation.
